# Determinants of practice location choices among physicians and medical students in Mali: insights into addressing medical deserts through evidence-based strategies

**DOI:** 10.1101/2025.02.25.25322851

**Authors:** Issa Kalossi, Dielika Coulibaly, Kassoum Alou N’diaye, Modibo Salia Drame, Djibril Sissoko, Thiery Almont, Kassoum Kayentao

## Abstract

The shortage of medical professionals in rural and remote areas is a global issue that significantly challenges equitable healthcare delivery. Worldwide, various studies have examined the motivations of medical professionals in choosing their practice location. However, for Mali, this topic remains underexplored, motivating us to conduct this study to identify factors influencing doctors’ workplace decisions in Mali. We conducted a cross-sectional study targeting doctors and final-year medical students. Using simple random sampling, we selected 358 respondents, 69% of whom were doctors. Approximately 38% of the respondents preferred rural areas for their practice, primarily citing career development and opportunities for continuing education (38%). The likelihood of choosing rural practice was higher (OR = 5.09; CI = [2.52-10.8]) among participants with family residing in rural areas. Conversely, financial incentives, access to technical platforms, and infrastructure favored urban practice. This study elucidates doctors’ motivations and identifies key factors associated with their choice of practice location.

## Introduction

A medical desert is characterized by a shortage of medical professionals in a specific geographical area. The lack of medical professionals in rural and remote areas is a global problem that poses significant challenges to equitable healthcare delivery. Both developed and developing countries report an uneven geographical distribution of medical professionals, favoring urban and affluent areas, despite the fact that rural communities experience greater health problems [1,2]. In 2012, more than half of the world’s population was living in rural areas, whereas less than a quarter of health professionals worked there [3]. This spatial disparity in the distribution of healthcare workers means that those who most need healthcare services receive the poorest care, reflecting Hart’s “Inverse Care Law” [4].

In Mali, community health centers (CSComs) represent the primary point of contact between the population and medical personnel. Nationally, only 32% of these centers have a physician, compared to 98% in Bamako, while the figure drops to just 29% in the rest of the country [5], this is well below the standards set by the World Health Organization. Additionally, the National Institute of Statistics estimates, in its latest report (2018), an average of 0.86 doctors per 10,000 inhabitants in rural regions versus 3.73 doctors per 10,000 inhabitants in Bamako [6].

To the best of our knowledge, no large-scale study has been conducted in Mali to gather medical professionals’ opinions on factors influencing their choice of practice location. Furthermore, this topic remains poorly documented in Mali’s unique context. This motivated us to investigate the main factors influencing the choice of practice location among doctors and final-year medical students in 2023. The findings of this study can inform targeted measures, public health actions, and workforce planning.

## Methods

### Study design

We conducted a cross-sectional study in Mali, surveying doctors and final-year medical students. Data were collected from May 1st to July 30th, 2023.

### Study population

Our study population included practicing doctors registered with the national medical council of Mali and final-year medical students. Not registered physicians and students who were not in their thesis year were excluded.

### Sampling method and sample size determination

We employed simple random sampling (without replacement) from the list of registered doctors and final-year medical students. To determine the minimum sample size (n), we used the following formula: [7]. 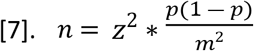 This calculation yielded a minimum required sample size of 347. Accounting 2% of the sample size as refusal rate, the final expected sample size was 354 participants.

### Data collection tools and procedures

Data were collected using an ad hoc questionnaire, tested prior to the study with students and physicians. It was inspired by prior studies on similar topics [8–16] and included cafeteria-style response options addressing the most frequently cited reasons during interviews. The questionnaire also captured sociodemographic and professional factors influencing health professionals’ practice location preferences. Designed in Microsoft Forms®, all key variables were mandatory to minimize missing data. The form link was distributed to selected participants via Short Message Service (SMS).

### Statistical analysis

The choice of practice location (urban or rural) served as the primary dependent variable, while other factors were treated as independent variables. A descriptive analysis was performed, with results presented in frequency tables. Statistical tests, including Pearson’s Chi-squared test and Fisher’s exact test, were used to compare proportions. To estimate odds ratios (ORs) and 95% confidence intervals (CIs) for each factor, univariate and multivariate logistic regression models were employed using all the key variables. Non-significant variables were removed from the final model using a stepwise approach based on the Akaike Information Criterion (AIC). A p-value ≤ 0.05 was considered statistically significant. Data analysis was performed using R software.

### Ethical considerations

This study received ethical approval from the National Ethics Committee for Health and Life Sciences (CNESS) in Mali, under reference number 23/04/MSDS/CNESS. To ensure the protection of respondents’ personal data, we used an anonymous questionnaire. Each participant was assigned an auto-generated number by the software in response order, preventing individual identification. As paper-written consent was not feasible for an online survey, respondents were informed about the objectives of the study and their voluntary participation. By completing and submitting the questionnaire, respondents were considered to have provided their informed consent to participate in the study.

## Results

At the end of the study, we collected 358 responses. The average age of the study population was 31 years (physicians = 34; students = 26), with a standard deviation of 6 years. For physicians, the average professional experience was 5 years, ranging from 0 to 32 years. Male respondents were predominant, accounting for 83.80% of participants, resulting in a male-to-female ratio of 5.17. Approximately 64.25% of respondents had urban family residences. Among the 358 respondents, 191 (53.35%) were born and/or raised in urban areas.

When asked about their preferred practice location, less than 38% (135/358) of respondents indicated a preference for working in rural areas if given the choice. Physicians constituted over 68% (246/358) of the study population, with nearly 77% of them working in the private sector. Bamako was the practice location for 51.22% of respondents, while the remaining 48.78% worked in other regions. More than 69% of respondents reported no prior experience working or interning in rural settings (Table 1).

**Table 1:** Sociodemographic and Professional Characteristics.

| Characteristics | Count (%) |
| --- | --- |
| **Sex** |  |
| Male/ Female | 300/58 (83,80) |
| **Family residence** |  |
| Urban/ Rural | 230/128 (64,25) |
| **Rural Origins** |  |
| Yes / No | 167/191 (46,65) |
| **Status** |  |
| Physician/ Student | 246/112 (68,72) |
| **Sector of employment** |  |
| Public sector / Private sector | 58/188 (23,58) |
| **Practice location** |  |
| Bamako (capital city)/Other regions | 126/120 (51,22) |
| **Rural experience** |  |
| Yes /No | 110/248 (30,73) |

Career development opportunities and access to continuing education were cited as the primary reasons for choosing a practice location by the majority of respondents (37.71%). This was followed by factors such as access to technical facilities or infrastructure. Financial reasons were the primary motivator for only 6.42% of respondents (Table 2).

**Table 2:** Distribution by Motivational Factors for Choosing a Practice Location.

| Motivational Factors | Reasons |  |
| --- | --- | --- |
|  | Primary Reason n (%) | Secondary Reason n (%) |
| Career development / Continuing education | 135 (37,71) | 81 (24,40) |
| Family/Personal factors | 99 (27,65) | 92 (27,71) |
| Technical facilities/Infrastructure | 51 (14,25) | 92 (27,71) |
| Accountability | 32 (8,94) | 27 (8,13) |
| Financial reasons | 23 (6,42) | 40 (12,05) |
| Other reasons | 18 (5,03) | - |

The intention to work in rural areas was higher among male respondents compared to female respondents, at 39% versus 31%, respectively; however, this difference was not statistically significant according to the Chi-squared test. Respondents with family residing in rural areas were significantly more likely to express a preference for rural practice compared to those without rural family ties, at 57.81% versus 26.52%. This difference was statistically significant (p < 0.001).

Similarly, rural practice intentions were statistically different for participants born or raised in rural areas compared to those who had only lived in urban settings. Participants with prior professional experience in rural settings, either through internships or work placements, were more likely to express an intention to work in rural areas compared to those without such experience (p < 0.001).

Regarding sociodemographic characteristics, neither univariate nor multivariate analyses showed a statistically significant association between gender and choice of practice location, with OR = 0.70 (CI = [0.38–1.27]) and OR = 0.67 (CI = [0.30–1.47]), respectively.

Respondents with family residing in rural areas were significantly more likely to choose rural practice in both univariate (OR = 3.80) and multivariate (OR = 5.09; p < 0.001) analyses. Rural origin was significantly associated with the choice of practice location in univariate analysis (OR = 2.54; p < 0.001), but this factor was not significant in multivariate analysis (OR = 0.87; p = 0.7).

Having rural experience was a factor favoring the intention to work in rural areas, with OR = 2.22 (CI = [1.36–3.70]) in univariate analysis and OR = 2.30 (CI = [0.77–6.81]) in multivariate analysis. Students were significantly more likely than practicing physicians to intend to work in rural areas, with an OR of 1.61 (CI = [1.02–2.54]) in univariate analysis and 2.05 (CI = [1.14–3.72]) in multivariate analysis (Table 3).

**Table 3:** Personal and Family Factors Associated with the Choice of Practice Location.

|  | Intention to Work |  | Univariate Analysis |  |  | Multivariate Analysis |  |  |
| --- | --- | --- | --- | --- | --- | --- | --- | --- |
| Factors | in Rural Areas n (%) | p value | OR | 95% CI | p value | OR | 95% CI | p value |
| Gender |  |  |  |  |  |  |  |  |
| Male | 117 (39,00) | 0,3 | 1,00 | — |  | 1,00 | — |  |
| Female | 18 (31,03) |  | 0,70 | 0,38 – 1,27 | 0,3 | 0,67 | 0,30 – 1,47 | 0,3 |
| Family residence |  |  |  |  |  |  |  |  |
| Urban | 61 (26,52) | <0,001 | 1,00 | — |  | 1,00 | — |  |
| Rural | 74 (57,81) |  | 3,80 | 2,41 – 6,03 | <0,001 | 5,09 | 2,52 – 10,8 | <0,001 |
| Rural origins |  |  |  |  |  |  |  |  |
| No | 44 (26,35) | <0,001 | 1,00 | — |  | 1,00 | — |  |
| Yes | 91 (47,64) |  | 2,54 | 1,64 – 4,00 | <0,001 | 0,87 | 0,41 – 1,78 | 0,7 |
| Status |  |  |  |  |  |  |  |  |
| Physician | 84 (34,15) | 0,039 | 1,00 | — |  | 1,00 | — |  |
| Student | 51 (45,54) |  | 1,61 | 1,02 – 2,54 | 0,04 | 2,05 | 1,14 – 3,72 | 0,017 |
| Rural experience |  |  |  |  |  |  |  |  |
| No | 107 (43,15) | 0,001 | 1,00 | — |  | 1,00 | — |  |
| Yes | 28 (25,45) |  | 2,22 | 1,36 – 3,70 | 0,002 | 2,30 | 0,77 – 6,81 | 0,13 |
| Note: OR: Odds Ratio; IC, Confidence Interval; p-value: * : <0.05, ** : <0.01, *** : <0.001 |  |  |  |  |  |  |  |  |

Note: OR: Odds Ratio; IC, Confidence Interval; p-value: \* : <0.05, \*\* : <0.01, \*\*\* : <0.001

(Table 4) For the main reasons influencing the choice of practice location among physicians and medical students, no statistically significant differences were found for participants citing personal or family-related reasons, as well as financial motivations.

**Table 4:**
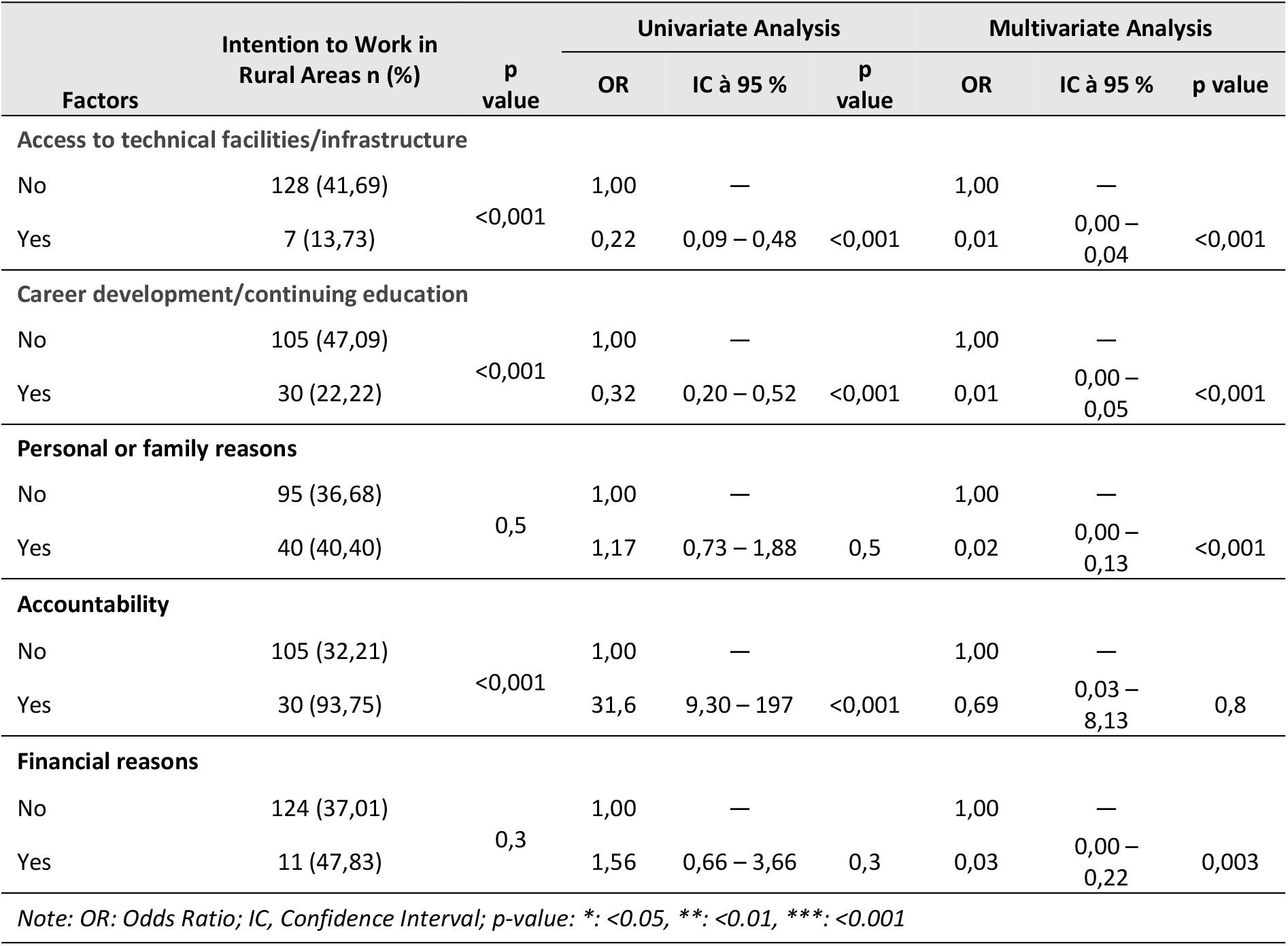
Professional Factors Influencing the Choice of Practice Location.

There was a statistically significant difference among participants who cited reasons such as accountability, access to technical facilities/infrastructure, and opportunities for career development or continuing education. Respondents who based their choice on access to technical facilities and infrastructure were less likely to intend to practice in rural areas (13.73%) compared to others. Among those motivated by career opportunities or continuing education, only 22.22% expressed an intention to work in rural areas. Conversely, 93.75% of those motivated by accountability chose rural areas as their preferred practice location. These differences were statistically significant (p < 0.001).

Regarding the motivations of medical personnel, choosing a location based on access to technical facilities was significantly associated with a lower likelihood of rural practice in both univariate analysis (OR = 0.22; CI = [0.09–0.48]) and multivariate analysis (OR = 0.01; CI = [0.00–0.04]). Motivations related to career development and continuing education were also negatively associated with rural practice intentions. This relationship was significant in both univariate analysis (OR = 0.32; CI = [0.20–0.52]) and multivariate analysis (OR = 0.01; CI = [0.00–0.05]).

There was no statistically significant relationship between motivations based on personal or family reasons and the choice of practice location in univariate analysis (OR = 1.17; CI = [0.73–1.88]). Participants motivated by personal or family factors were less likely to choose rural practice (OR = 0.02; p < 0.001).

Respondents motivated by accountability were significantly more likely to intend to work in rural areas (OR = 31.6; CI = [9.30–197]) in univariate analysis. However, this relationship was not statistically significant in multivariate analysis (OR = 0.69; CI = [0.03–8.13]).

Financial motivation was not a statistically significant factor in the choice of practice location in multivariate analysis. However, it was negatively associated with rural practice intentions (OR = 0.03; CI = [0.00–0.22]).

**Figure 1:**
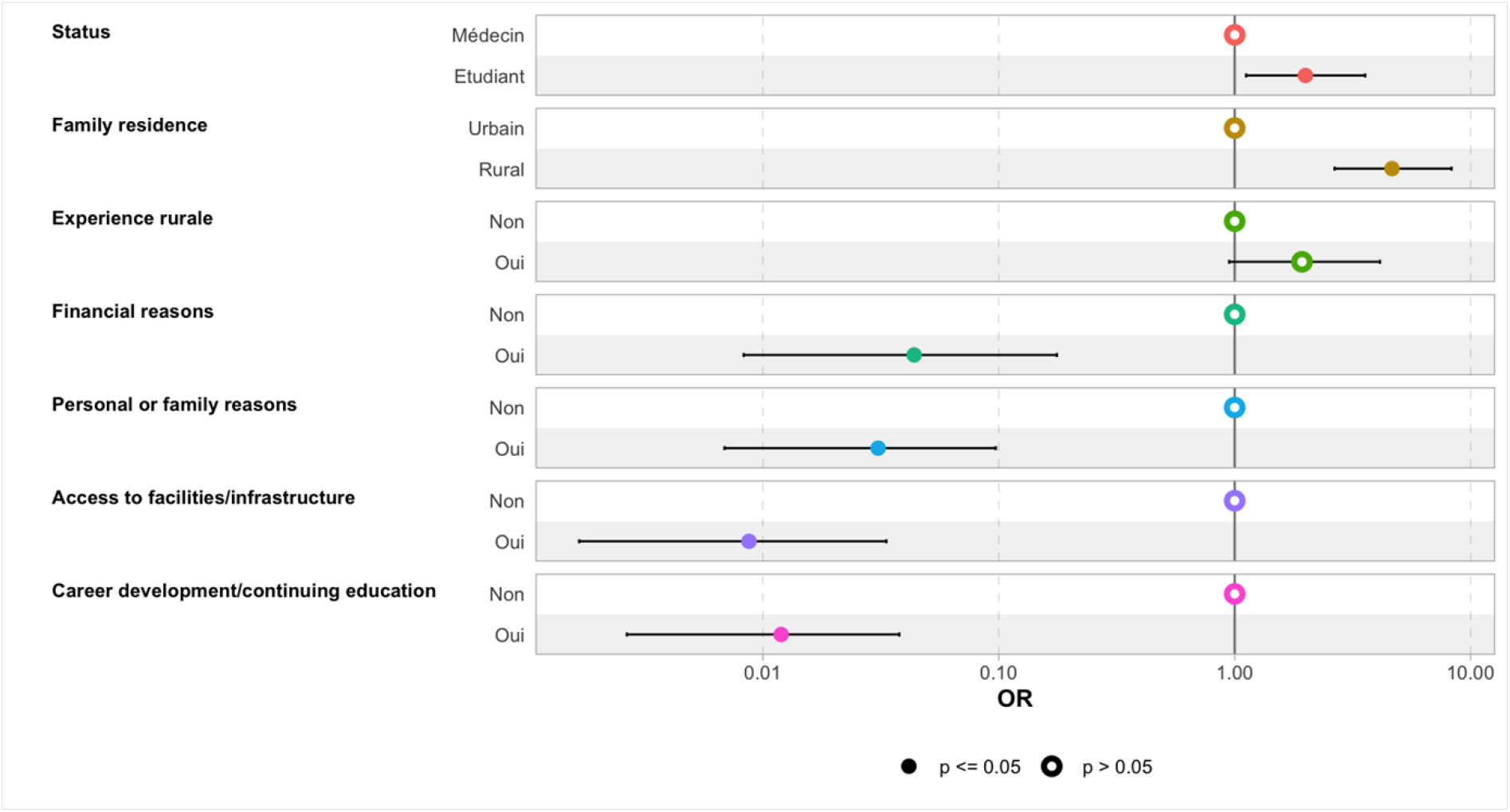
Final Multivariate Regression Model of Associated Factors.

According to the final model (Figure 3), the family residence of the respondents had a significant influence on their choice of practice location. Students were also more likely to choose rural areas compared to practicing physicians. Having prior experience in rural settings was another factor positively influencing the choice of rural practice.

Conversely, financial motivation, personal or family reasons, career opportunities, continuing education, and access to technical facilities were all significantly and negatively associated with the decision to practice in rural areas.

## Recommendations and Proposed Solutions

To promote an equitable distribution of physicians across the country, our results have highlighted the following key approaches:

### Recruitment

Terms such as “recruitment,” “direct integration,” or “massive recruitment” were frequently mentioned by respondents. In Mali, recruitment into the state public service is conducted through competitive examinations. Out of an average of 350 physicians trained annually, only about twenty are integrated into the public service each year. This limited recruitment may contribute to the shortage of physicians in certain remote or underserved areas. In the absence of large-scale national recruitment, some respondents suggested community-based recruitment strategies, explicitly specifying the practice location, to address geographic disparities.

### Decentralization

The decentralization of technical facilities and improved access to healthcare infrastructure were identified by some respondents as factors that could encourage physicians to settle in rural areas. The decentralization of continuing medical education was also proposed as an incentive. Currently, in Mali, access to general medical education and medical specialization is only available in Bamako (the capital city).

### Financial Incentives and Benefits

Financial motivation through increased salaries, special allowances for physicians working in rural areas, and other incentive-based benefits were suggested by several respondents as potential solutions to reduce medical deserts.

### Political Will / Leadership

The lack of political commitment in the health sector was also highlighted. Some participants stated that strong political will is essential to address the issue of medical deserts effectively.

### Rural Internships

The establishment of a mandatory rural internship program for final-year medical students was proposed. This measure could enable early-career physicians to acquire foundational knowledge and practical skills related to rural medical practice.

## Discussion

To the best of our knowledge, this study, which aimed to analyze the factors associated with the choice of practice location among physicians and final-year medical students, is the first of its kind in our context. It provided insights into the motivations and influencing factors behind the practice location preferences of physicians and medical students.

### Family Residence

The results revealed a strong association between family residence and the respondents’ choice of practice location. Having family in rural areas positively influenced the decision to practice in rural settings. Among the 128 respondents whose families resided in rural areas, approximately 58% chose rural practice, a statistically significant difference. This factor was also associated with the choice of practice location in univariate (OR = 3.80) and multivariate analyses (OR = 5.09). This finding aligns with other studies conducted in Botswana[17], Ethiopia [18] and Nepal [19]. However, other research, such as that by Liu et al. [20], de Playford et al. [21] et de Grobler et al. [22] found family residence to be less influential..

### Rural Experience

A statistically significant relationship was observed between rural experience and the choice of practice location in univariate analysis. Respondents with rural experience were 2.22 times (crude OR) more likely to intend to practice in rural areas than those without such experience. However, this difference was not significant in multivariate analysis (CI = [0.77–6.81]). This result is consistent with findings from other countries [21,23,24].

### Status

Our multivariate logistic regression model also identified status as a significant factor influencing respondents’ decisions. Among the 112 students in our sample, nearly 46% expressed an intention to practice in rural areas, compared to 34% of practicing physicians. This difference was statistically significant according to the Chi-squared test. In multivariate regression, students were 2.05 times more likely than physicians to intend to work in rural areas (CI = [1.14–3.72]). This finding aligns with those of Raul et al. [25], who reported a strong rural practice intention among 21% of students.

### Financial Motivation

Our study found that financial motivation was negatively associated with the decision to practice in rural areas. Physicians and students driven by financial incentives were more likely to prefer urban practice, with an OR of 0.03 (CI = [0.00–0.22]) in multivariate analysis. This result is comparable to studies conducted in Kenya [26] and Ghana [27], where financial motivation was a key factor influencing respondents’ choices.

### Personal or Family Reasons

Personal or family reasons were also associated with the respondents’ choice of practice location in multivariate analysis. However, contrary to studies in Tanzania [28] and a 2016 systematic review [29], our findings revealed that participants citing personal or family reasons were more likely to prefer urban practice. This factor was statistically significant after adjustment (OR = 0.02; CI = [0.00–0.13]).

### Access to Infrastructure and Technical Facilities

Access to infrastructure and technical facilities also emerged as a motivational factor. Approximately 14% of respondents cited this as their primary reason for choosing a practice location, with nearly 28% mentioning it as a secondary reason. Over 86% of respondents motivated by this factor preferred urban practice. This preference was significant in both univariate (OR = 0.22) and multivariate analyses (OR = 0.01; CI = [0.00–0.04]). Our result was comparable to a study conducted in Kenya [26] and Ghana [27], where inadequate access to electricity, equipment, and transport was found to be a determining factor in the choices made by the respondents. The lack of housing, inadequate remuneration for health staff, and the poor condition of healthcare facilities contribute to an unfavorable work environment. Inadequate working conditions, combined with low job satisfaction and stability, were cited as factors that could demotivate healthcare workers and impact their decision to practice in rural areas, according to the study by Dielemann et al. [11] and Willis-Schattuck et al. [30]. Other authors have also reported that housing is one of the factors that attract and retain rural doctors [31]. Hospital accommodation is often the only realistic option for rural doctors because many hospitals are in extremely underdeveloped areas, with inadequate housing available in communities outside the hospital.

### Career Development and Continuing Education

Another significant motivation influencing the choice of practice location among respondents was the opportunity for career advancement and continuing education. In our study, 135 respondents (38%) stated that career progression and continuing education opportunities were the primary reasons for their decision, while 24% cited these as secondary reasons. Among participants whose primary reason was career development or continuing education, 78% opted for urban practice, a statistically significant difference based on the Chi-squared test (p < 0.001).

Respondents who prioritized career development or continuing education were more likely to choose urban settings as their practice location, as shown by univariate logistic regression (OR = 0.32; CI = [0.20–0.52]) and multivariate regression (adjusted OR = 0.01; CI = [0.00–0.04]). These findings align with those of a systematic review [30], which indicated that health professionals were reluctant to work in rural areas due to fewer career development opportunities compared to urban settings [32]. Studies have shown that health professionals feel proud and motivated when they perceive opportunities for progression.

### Accountability

Accountability was another factor described in the literature as significantly influencing the decision of health professionals to practice in rural areas. According to our study, accountability was cited as the primary reason by approximately 9% of respondents. Among these, nearly 94% expressed an intention to work in rural areas if given the choice, a statistically significant difference based on the Chi-squared test.

In logistic regression analysis, participants motivated by accountability were 32 times more likely (crude OR) to intend to work in rural areas compared to those citing other reasons. However, this difference was not statistically significant after adjustment for all other variables (OR = 0.69; CI = [0.03– 8.13]). Some studies have similarly reported that feelings of belonging, and accountability are key determinants influencing participants’ decisions to remain in rural areas [33].

### Rural Origins

In our study, participants born or raised in rural areas accounted for approximately 47% of respondents. Among them, nearly 58% expressed an intention to practice in rural areas, a statistically significant difference based on the Chi-squared test (p < 0.001). Respondents with rural origins were 2.54 times more likely (crude OR) to choose rural practice compared to those with urban origins. However, this trend was not statistically significant after adjustment in multivariate logistic regression.

In contrast to our findings, other studies have reported statistically significant relationships between rural origin and the choice to practice in rural areas. These studies found that rural origin was positively associated with the preference for rural practice locations [21,23,24].

### Gender Imbalance

We also evaluated the influence of other sociodemographic characteristics, such as gender, on respondents’ choice of practice location. Female respondents were underrepresented in our study, accounting for only 16% of participants. Among female respondents, 31% expressed an intention to work in rural areas compared to 39% of male respondents. However, this difference was not statistically significant (p = 0.3).

In both univariate and multivariate analyses, female respondents were more inclined to choose urban settings. The ORs were 0.70 (CI = [0.38–1.27]) and 0.67 (CI = [0.30–1.47]), respectively.

### Strengths and Limitations

One of the key strengths of this study lies in its originality, particularly the inclusion of final-year medical students as part of the study population. This group is rarely investigated in similar contexts, especially in Mali, despite representing the future workforce whose preferences could significantly shape the healthcare landscape in the coming years. By capturing the perspectives of both practicing physicians and students, our study offers a more comprehensive understanding of the determinants influencing practice location choices.

However, some limitations must be acknowledged. In fact, the expressed intentions of medical students regarding their future practice location may not necessarily translate into actual professional choices later in their careers. The existing literature suggests that career intentions often evolve under the influence of personal, professional, and contextual factors. Also, the geographic scope of the study may limit the generalizability of the findings to other regions beyond the surveyed areas.

These considerations reflect our commitment to a critical and reflexive interpretation of the results, ensuring that conclusions are drawn with appropriate caution.

## Conclusion

Medical deserts are a phenomenon affecting all countries worldwide, both developed and developing. This issue has serious consequences for public health, particularly in rural and remote populations.

This study provided insights into the motivations and influencing factors behind physicians’ and medical students’ choices of practice location. Factors such as family residence, rural experience, and professional motivations were significantly associated with respondents’ decisions regarding their practice location.

These factors could serve as levers for ensuring the equitable distribution of physicians across the national territory.

## Data Availability

All data underlying the findings of this study are fully available without restriction.

## References

1. Wakerman J, Humphreys JS. Rural health: why it matters. Med J Aust [Internet]. 2002 [cité 25 avr 2022];176(10):457–8. Disponible sur: https://onlinelibrary.wiley.com/doi/abs/10.5694/j.1326-5377.2002.tb04514.x

2. Frélaut M. Les déserts médicaux. Regards [Internet]. 2018 [cité 16 janv 2023];53(1):105–16. Disponible sur: https://www.cairn.info/revue-regards-2018-1-page-105.htm

3. Carmen R Mandy, Kolstad, Julie R, Rockers, Peter C, Dolea. How to conduct a discrete choice experiment for health workforce recruitment and retention in remote and rural areas : a user guide with case studies [Internet]. World Bank. [cité 15 févr 2025]. Disponible sur: https://documents.worldbank.org/en/publication/documents-reports/documentdetail/en/586321468156869931

4. Hart JT. THE INVERSE CARE LAW. The Lancet [Internet]. 27 févr 1971 [cité 25 avr 2022];297(7696):405–12. Disponible sur: https://www.thelancet.com/journals/lancet/article/PIIS0140-6736(71)92410-X/fulltext

5. Évaluation du Système de Santé au Mali | HFG [Internet]. 2017 [cité 25 mars 2022]. Disponible sur: https://www.hfgproject.org/evaluation-du-systeme-de-sante-au-mali/

6. INSTAT. ANNUAIRE STATISTIQUE DU MALI [Internet]. Institut National de la Statistique du Mali|INSTAT. [cité 28 mars 2022]. Disponible sur: https://www.instat-mali.org/fr/publications/annuaire-statistique-du-mali-

7. Maitre Y. Méthode d’échantillonnage dans les études épidémiologiques transversales nationales auprès des professionnels d santé en France application odontologie. 2021;

8. Helmuth. Influences on the choice of health professionals to practise in rural areas. 2007;2.

9. McDonald F, Simpson C. Challenges for rural communities in recruiting and retaining physicians. Can Fam Physician [Internet]. sept 2013 [cité 13 mars 2022];59(9):915–7. Disponible sur: https://www.ncbi.nlm.nih.gov/pmc/articles/PMC3771713/

10. Lucas-Gabrielli V, Chevillard G. « Déserts médicaux » et accessibilité aux soins : de quoi parle-t-on ? médecine/sciences [Internet]. juin 2018 [cité 13 mars 2022];34(6-7):599–603. Disponible sur: https://www.medecinesciences.org/10.1051/medsci/20183406022

11. Dieleman M, Cuong PV, Anh LV, Martineau T. Identifying factors for job motivation of rural health workers in North Viet Nam. Hum Resour Health [Internet]. 5 nov 2003 [cité 10 avr 2022];1(1):10. Disponible sur: 10.1186/1478-4491-1-10

12. Hancock C, Steinbach A, Nesbitt TS, Adler SR, Auerswald CL. Why doctors choose small towns: A developmental model of rural physician recruitment and retention. Soc Sci Med [Internet]. nov 2009 [cité 13 mars 2022];69(9):1368–76. Disponible sur: https://linkinghub.elsevier.com/retrieve/pii/S0277953609005152

13. Brooks RG, Walsh M, Mardon RE, Lewis M, Clawson A. The Roles of Nature and Nurture in the Recruitment and Retention of Primary Care Physicians in Rural Areas: A Review of the Literature. Acad Med [Internet]. août 2002 [cité 14 mars 2022];77(8):790–8. Disponible sur: http://journals.lww.com/00001888-200208000-00008

14. Henderson RW. Staffing rural hospitals. Strategies for survival. Can Fam Physician [Internet]. juin 1996 [cité 14 mars 2022];42:1057–72. Disponible sur: https://www.ncbi.nlm.nih.gov/pmc/articles/PMC2146484/

15. Missinne S, Luyten S. Les médecins généralistes en région bruxelloise : qui sont-ils, où pratiquent-ils et où se situent les potentielles pénuries? [Internet]. Bruxelles: Commission communautaire française-Cocof; 2018. 43 p. Disponible sur: http://www.ccc-ggc.brussels/fr/observatbru/publications/dossier-20182-les-medecins-generalistes-en-region-bruxelloise-qui-sont-ils

16. Lehmann U, Dieleman M, Martineau T. Staffing remote rural areas in middle- and low-income countries: a literature review of attraction and retention. BMC Health Serv Res. 23 janv 2008;8:19.

17. Arscott-Mills T, Kebaabetswe P, Tawana G, Mbuka DO, Makgabana-Dintwa O, Sebina K, et al. Rural exposure during medical education and student preference for future practice location - a case of Botswana. Afr J Prim Health Care Fam Med [Internet]. 10 juin 2016 [cité 18 juill 2023];8(1):1039. Disponible sur: https://www.ncbi.nlm.nih.gov/pmc/articles/PMC4926713/

18. Deressa W, Azazh A. Attitudes of undergraduate medical students of Addis Ababa University towards medical practice and migration, Ethiopia. BMC Med Educ [Internet]. 6 août 2012 [cité 18 juill 2023];12(1):68. Disponible sur: 10.1186/1472-6920-12-68

19. Sapkota BP, Amatya A. What factors influence the choice of urban or rural location for future practice of Nepalese medical students? A cross-sectional descriptive study. Hum Resour Health [Internet]. 10 nov 2015 [cité 18 juill 2023];13(1):84. Disponible sur: 10.1186/s12960-015-0084-5

20. Liu J, Zhu B, Zhang N, He R, Mao Y. Are Medical Graduates’ Job Choices for Rural Practice Consistent with their Initial Intentions? A Cross-Sectional Survey in Western China. Int J Environ Res Public Health [Internet]. janv 2019 [cité 26 janv 2023];16(18):3381. Disponible sur: https://www.mdpi.com/1660-4601/16/18/3381

21. Playford D, Ngo H, Gupta S, Puddey IB. Opting for rural practice: the influence of medical student origin, intention and immersion experience. Med J Aust [Internet]. août 2017 [cité 18 juill 2023];207(4):154–8. Disponible sur: https://onlinelibrary.wiley.com/doi/abs/10.5694/mja16.01322

22. Grobler L, Marais BJ, Mabunda SA, Marindi PN, Reuter H, Volmink J. Interventions for increasing the proportion of health professionals practising in rural and other underserved areas. Cochrane Database Syst Rev [Internet]. 2009 [cité 18 juill 2023];(1). Disponible sur: https://www.cochranelibrary.com/cdsr/doi/10.1002/14651858.CD005314.pub2/full

23. Gt S, Rj S. Nature or nurture: the effect of undergraduate rural clinical rotations on pre-existent rural career choice likelihood as measured by the SOMERS Index. Aust J Rural Health [Internet]. avr 2012 [cité 18 juill 2023];20(2). Disponible sur: https://pubmed.ncbi.nlm.nih.gov/22435768/

24. Puddey IB, Mercer A, Playford DE, Riley GJ. Medical student selection criteria and socio-demographic factors as predictors of ultimately working rurally after graduation. BMC Med Educ [Internet]. 14 avr 2015 [cité 18 juill 2023];15(1):74. Disponible sur: 10.1186/s12909-015-0359-5

25. Borracci RA, Arribalzaga EB, Couto JL, Dvorkin M, Ahuad Guerrero RA, Fernandez C, et al. Factors affecting willingness to practice medicine in underserved areas: a survey of Argentine medical students. Rural Remote Health. 2015;15(4):3485.

26. Ojakaa D, Olango S, Jarvis J. Factors affecting motivation and retention of primary health care workers in three disparate regions in Kenya. Hum Resour Health. 6 juin 2014;12:33.

27. Johnson JC, Nakua E, Dzodzomenyo M, Agyei-Baffour P, Gyakobo M, Asabir K, et al. For money or service?: a cross-sectional survey of preference for financial versus non-financial rural practice characteristics among Ghanaian medical students. BMC Health Serv Res. 3 nov 2011;11:300.

28. Shemdoe A, Mbaruku G, Dillip A, Bradley S, William J, Wason D, et al. Explaining retention of healthcare workers in Tanzania: moving on, coming to « look, see and go », or stay? Hum Resour Health. 19 janv 2016;14:2.

29. Budhathoki SS, Zwanikken PAC, Pokharel PK, Scherpbier AJ. Factors influencing medical students’ motivation to practise in rural areas in low-income and middle-income countries: a systematic review. BMJ Open. 22 févr 2017;7(2):e013501.

30. Willis-Shattuck M, Bidwell P, Thomas S, Wyness L, Blaauw D, Ditlopo P. Motivation and retention of health workers in developing countries: a systematic review. BMC Health Serv Res [Internet]. 4 déc 2008 [cité 19 juill 2023];8(1):247. Disponible sur: 10.1186/1472-6963-8-247

31. Villiers MD, Villiers PD. Doctors’ views of working conditions in rural hospitals in the Western Cape. South Afr Fam Pract [Internet]. 24 juin 2004 [cité 22 juill 2023];46(3):21–6. Disponible sur: https://www.ajol.info/index.php/safp/article/view/13100

32. Kotzee TJ, Couper ID. What interventions do South African qualified doctors think will retain them in rural hospitals of the Limpopo province of South Africa? Rural Remote Health. 2006;6(3):581.

33. Sheikh K, Rajkumari B, Jain K, Rao K, Patanwar P, Gupta G, et al. Location and vocation: why some government doctors stay on in rural Chhattisgarh, India. Int Health [Internet]. 1 sept 2012 [cité 28 juill 2023];4(3):192–9. Disponible sur: 10.1016/j.inhe.2012.03.004

